# Using genetic information to define idiopathic pulmonary fibrosis in UK Biobank

**DOI:** 10.1101/2022.04.01.22273306

**Authors:** Olivia C Leavy, Richard J Allen, Luke M Kraven, Ann Morgan, Martin D Tobin, Jennifer K Quint, R Gisli Jenkins, Louise V Wain

## Abstract

**Introduction:** Idiopathic pulmonary fibrosis (IPF) is a rare lung disease characterised by progressive scarring in the alveoli. IPF can be defined in population studies using electronic healthcare records (EHR) but recent genetic studies of IPF using EHR have shown an attenuation of effect size for known genetic risk factors when compared to clinically-derived datasets, suggesting misclassification of cases.

**Methods:** We used EHR (ICD-10, Read (2 & 3)) and questionnaire data to define IPF cases in UK Biobank, and evaluated these definitions using association results for the largest genetic risk variant for IPF (rs35705950-T, *MUC5B*). We further evaluated the impact of exclusions based on co-occurring codes for non-IPF pulmonary fibrosis and restricting codes according to changes in diagnostic practice.

**Results:** Odds ratio (OR) estimates for rs35705950-T associations with IPF defined using EHR and questionnaire data in UK Biobank were significant and ranged from 2.06 to 3.09 which was lower than those reported using clinically-derived IPF datasets (95% confidence intervals: 3.74, 6.66). Code-based exclusions of cases gave slightly closer effect estimates to those previously reported, but sample sizes were substantially reduced.

**Discussion:** We show that none of the UK Biobank IPF codes replicate the effect size for the association of rs35705950-T on IPF risk when using clinically-derived IPF datasets. Further code-based exclusions also did not lead to effect estimates closer to those expected. Whilst the apparent increased sample sizes available for IPF from general population cohorts may be of benefit, future studies should take these limitations of the case definition into account.

**Key Messages:** *What is already known on this topic:* UK Biobank is a very large prospective cohort that can be utilised to increase sample sizes for studies of rare diseases such as idiopathic pulmonary fibrosis (IPF). However, effect size estimates for genetic risk factors for IPF in UK Biobank and other general population cohorts, when defining cases using electronic healthcare records (EHR), are smaller than those estimated from clinically-derived IPF datasets.

*What this study adds:* Using Hospital Episode Statistics (HES) data, primary care data, death registry data and self-report data in UK Biobank, we used the association rs35705950-T, the largest genetic risk factor for IPF, to evaluate code-based definitions of IPF. We show that none of the available IPF coding replicates the effect size for rs35705950-T on IPF risk that is observed in clinically-derived IPF datasets.

*How this study might affect research, practice or policy:* Research using large general population cohorts and datasets for observational studies of IPF should take these limitations of EHR definitions of IPF into consideration.

## Introduction

Interstitial lung diseases (ILD) are a large group of pulmonary conditions that are characterised by inflammation and scarring of the alveoli. Idiopathic Pulmonary Fibrosis (IPF) is the most common ILD in European ancestry populations and the most well studied genetically. IPF has a prevalence of 50 per 100,000 and incidence of 6,000 new cases per year in the UK (1). Median survival time with IPF is 3 to 5 years, which is a worse prognosis than many common cancers (2).

Genome-wide association studies have identified 20 independent single nucleotide polymorphisms (SNPs) associated with IPF risk to date (3-9). Of these, the biggest genetic risk factor for IPF is a single nucleotide polymorphism (SNP) in the promoter region of the *MUC5B* gene (rs35705950), involved in regulating mucus production. This SNP has an unusually large effect on risk for a common variant. Each copy of the risk allele T is associated with a 4-5 times increased risk of IPF (7, 10) and may explain more variation in disease liability than the other common IPF susceptibility SNPs combined (11).

Studies aiming to understand the pathophysiology of disease often depend on the availability of linked clinical and bio-sample data, for example, for genetic and biomarker studies. IPF is an uncommon disease and most such datasets for IPF have been developed from dedicated IPF cohort studies, registries and clinical trials, which are usually modest in size consisting of only a few hundred cases. Very large general population cohorts, such as UK Biobank, which has extensive linked molecular and phenotypic data, represent a valuable resource for increasing IPF case sample sizes, and hence statistical power for molecular epidemiological studies including genetic studies.

The observed effect size estimates for rs35705950 on IPF risk in UK Biobank and other general population cohorts, when defining cases using the ICD-10 J84.1 code (Other interstitial pulmonary diseases with fibrosis), are smaller than those estimated in clinically-derived datasets (12). Whilst this attenuation may be due to some misclassification of IPF cases, this may to some extent be mitigated by the substantial gain in statistical power that can be leveraged from very large biobanks. However, more accurate classification of cases and controls in biobanks could provide more accurate effect estimates for use in further analyses.

Given this, we proposed that the IPF susceptibility association effect size of rs35705950 (*MUC5B* promoter SNP), the largest and most consistently reported genetic risk variant for IPF, could be used to evaluate the choice of codes to define IPF cases and we applied this approach in UK Biobank.

To our knowledge, this is the first time that a genetic effect has been used to assess the way IPF is currently being defined in the UK Biobank. Our findings have wide applicability for how IPF is defined in general population cohorts with linked hospital data for a range of different study types.

## Methods

### Data

UK Biobank is a large-scale prospective cohort study containing over 500,000 volunteers recruited in the UK from 2006 to 2010 at ages 49-69 years (13, 14). UK Biobank has baseline, genetic and linked health-related outcome data that includes Hospital Episode Statistics (HES) data, primary care data and death registry data.

The ICD-10 code J84.1 was initially used to define IPF in HES (2020 release, last admission date for whole UK Biobank: 30-06-2020) and mortality (May 2020 version, last date of death for whole UK Biobank: 22-05-2020) data in UK Biobank. HES and mortality data were available for the whole UK Biobank cohort.

Two self-reported pulmonary fibrosis variables were available for the UK Biobank cohort. At baseline assessment, participants were asked by a trained nurse to self-report any non-cancer illnesses (field id 20002) which included ‘pulmonary fibrosis’. During an online follow-up survey about work environment conducted in 2015, 121,270 participants answered a question about whether a doctor had ever diagnosed them with IPF (field id: 22135, version July 2017).

Primary care data were available for 230,105 participants (last event recorded by General Practitioner: 18-08-2019). Eight primary care codes, a mixture of Read 2 and Read 3 codes, were used to define IPF (15). We also extracted codes for prescriptions of the two anti-fibrotic treatments that are currently only licensed for use by IPF patients (Nintedanib and Pirfenidone).

All codes used to define IPF cases in UK Biobank are given in **Table 1**. UK Biobank participants who self-reported as having IPF or pulmonary fibrosis, or had any one of the codes in HES, death registrar or primary care were combined together and defined as the “IPF subset”. Controls were defined as individuals who had linked primary care data, but had not been defined as an IPF case in any of the data sources. We further matched controls to cases for age at admission, sex, ever-smoker status and genetically determined ethnicity (50,924 individuals) (described in **Supplementary Methods**).

**Table 1:** List of codes used to define IPF in UK Biobank.

|  | HES Definition | Mortality definition | Primary care definition | Self-reported IPF | Self-reported pulmonary fibrosis | IPF |
| --- | --- | --- | --- | --- | --- | --- |
| **Data availability** | Linked to everyone in UK Biobank | Linked to everyone in UK Biobank | Available for 230,105 participants | Available for 121,270 participants | Asked of all participants at baseline | - |
| **Number of IPF cases** | 2,303 | 580 | 442 | 107 | 142 | 2,629 |
| **Definition** | J84.1 ICD-10 code used in HES data | J84.1 ICD-10 code used in data fields:<br>- 40001 (Underlying cause of death)<br>- 40002 (Contributory causes of death) | Any of following Read 2 or 3 codes appears in primary care data:<br>- H563./XE0Yb: Idiopathic fibrosing alveolitis<br>- H563./X102v: Hamman-Rich syndrome<br>- H563./XE0Yb: Cryptogenic fibrosing alveolitis<br>- H563./XE0Yb: Idiopathic pulmonary fibrosis<br>- H5631/H5631: Diffuse pulmonary fibrosis<br>- H5633/X102v: Usual interstitial pneumonitis<br>- H563z/H563z: Idiopathic fibrosing alveolitis not otherwise specified<br>- H5632/X102u: Pulmonary fibrosis<br>And/or has prescription for:<br>- Pirfenidone<br>- Nintedanib | Answered “idiopathic pulmonary fibrosis” to question “Has a doctor ever told you that you have had any of the conditions below?” (data field 22135) | Answered “pulmonary fibrosis” for “Non-cancer illness code, self-reported” (data field 20002) | Defined as an IPF case in any of the other IPF definitions |

The *MUC5B* promotor SNP rs35705950 was directly genotyped using either the Affymetrix UK BiLEVE Axiom array or the Affymetrix UK Biobank array. We excluded individuals who failed genome-wide genotype quality control and those with missing ethnicity (**Supplementary Methods**).

### Analyses

We tested the association of rs35705950 with IPF risk in UK Biobank using a logistic regression model and adjusted for the first ten genetic principal components. As the frequency of the *MUC5B* SNP varies considerably between populations, we limited genetic analyses to individuals who were of European ancestry. We compared the effect size (odds ratio, OR) of the association using each IPF definition, with that reported in the largest GWAS of IPF susceptibility conducted to date (9), as well as meta-analysed ORs given in a published *MUC5B* meta-analysis (10). The IPF cases included in Allen *et al* (9) were diagnosed as cases using American Thoracic Society and European Respiratory Society guidelines (16). For the Zhu *et al MUC5B* meta-analysis they included six studies investigating the association between rs35705950 and IPF in the Caucasian population published between 2011 and 2015. We considered these previously reported rs35705950 IPF susceptibility effect size (odds ratio) as the ‘gold standard’ against which to evaluate codes for IPF in UK Biobank.

Using the ICD-10 (HES and mortality) defined dataset, we evaluated the effect of excluding participants with co-occurring ICD-10 codes (in HES or mortality) that might indicate misclassification. We then repeated the association testing for the *MUC5B* SNP and compared the effect size to the ‘gold standard’. Specifically, we excluded the following categories of codes:

- codes for non-IPF medical conditions that cause pulmonary fibrosis (codes defined and collated by Bellou *et al* (17) (**Supplementary Table 1**)
- J84.1 ICD-10 code occurrence before the year that the most recent clinical guidelines for diagnosis of IPF (16) was published (2018)

## Results

### IPF definitions in UK Biobank

Of those with non-missing ancestry, there were 2,629 individuals with one or more codes indicative of IPF (IPF subset) **(Table 1)**. Of the 2,629 participants, 1,240 had linked primary care records and 432 had answered data field 22135 (self-reported IPF).

The eight individuals with an IPF medication code also had a primary care IPF Read 2 or Read 3 code indicative of IPF and a J84.1 ICD-10 code in HES. Five of the eight with a medication code also had a mortality J84.1 ICD-10 code and two self-reported as having IPF.

There were 741 participants (28% of IPF cases) who had codes indicative of IPF from at least two different sources and 181 participants (7% of IPF cases) who had codes indicative of IPF from at least three different sources. There were 202 individuals that had data available from four sources and who were defined as an IPF case in at least one of the four sources, excluding mortality (i.e. had linked HES and primary care data, and had responded positively or negatively to both of the two self-report questions) (**Supplementary Figure 1**). Of these, five had a code indicative of IPF from all four sources.

### Association of MUC5B SNP rs35705950 with IPF risk

In UK Biobank, the *MUC5B* SNP rs35705950 was genome-wide significantly associated with IPF risk (*p*-value <5×10^−8^) for all but the self-reported pulmonary fibrosis definition (*p*-value=1.00×10^−6^) **(Figure 1 & Supplementary Table 2)**. The observed ORs ranged from 2.06-3.09 and were all lower than the OR and below the confidence intervals reported in Allen *et al* 2021 (OR = 5.06, 95% CI: 4.69, 5.47) and the published *MUC5B* meta-analysis (Zhu *et al* 2015 (10): 4.99 [3.74, 6.66]). Self-reported IPF cases had the closest OR to previously published estimates (OR=3.09, 95% CI = 2.28, 4.18) but was the smallest case sample size. Defining IPF using the “J84.1” ICD-10 code in HES data, or the self-reported pulmonary fibrosis, gave the OR furthest away from previously reported estimates but HES had the largest case sample sizes.

**Figure 1:**
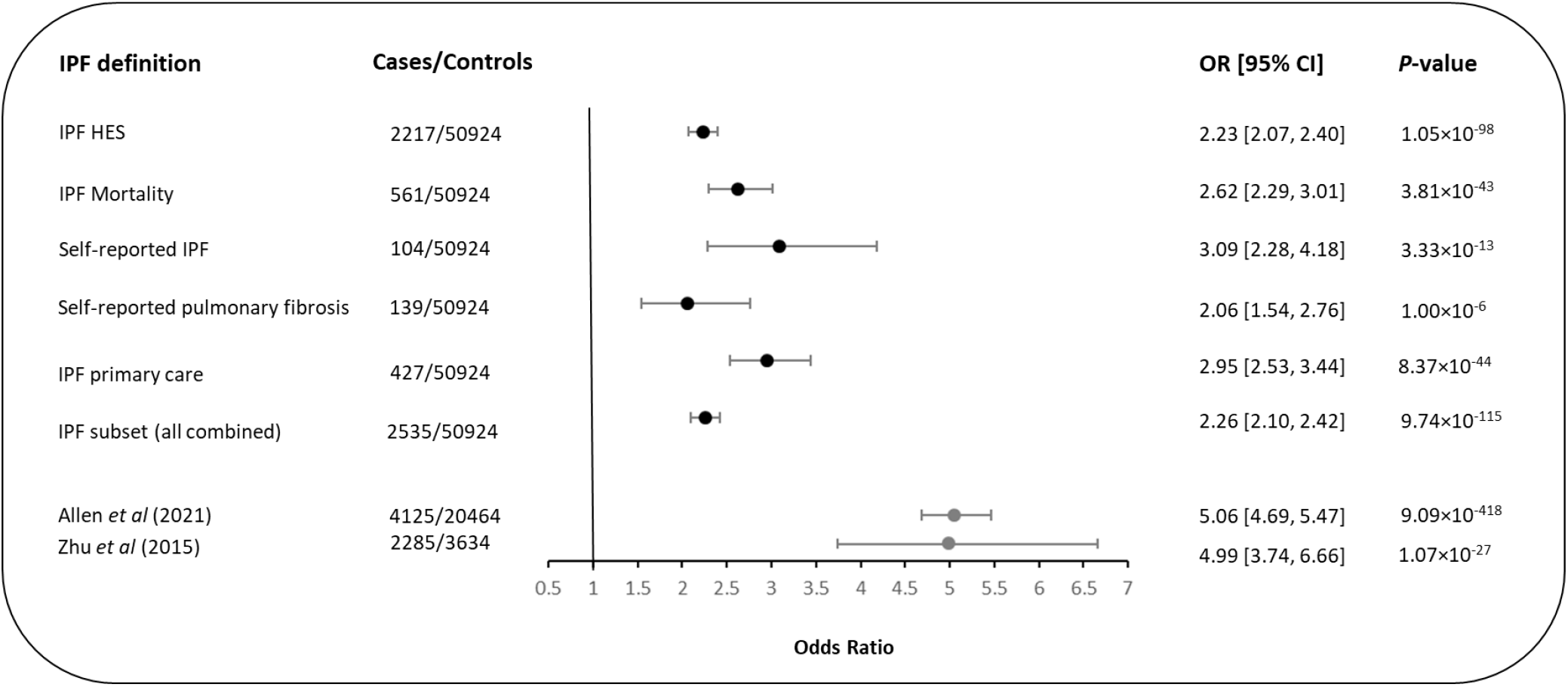
Effect size estimates of rs35705950 T allele association with IPF risk using different IPF case definitions. Each line shows the effect size estimate and confidence interval for the association between rs35705950 and IPF risk using the different methods for defining IPF in UK Biobank. Estimates in grey are the reference effect size estimates taken from Allen *et al* (2021) and Zhu *et al* (2015).

### Refining IPF case definition

Removing cases with a co-occurring code indicative of being non-IPF ILD or removing cases defined by the occurrence of a J84.1 code before January 2018, led to slightly closer effect estimates to those previously reported, but with substantially reduced sample sizes (**Figure 2, Supplementary Table 3**).

**Figure 2:**
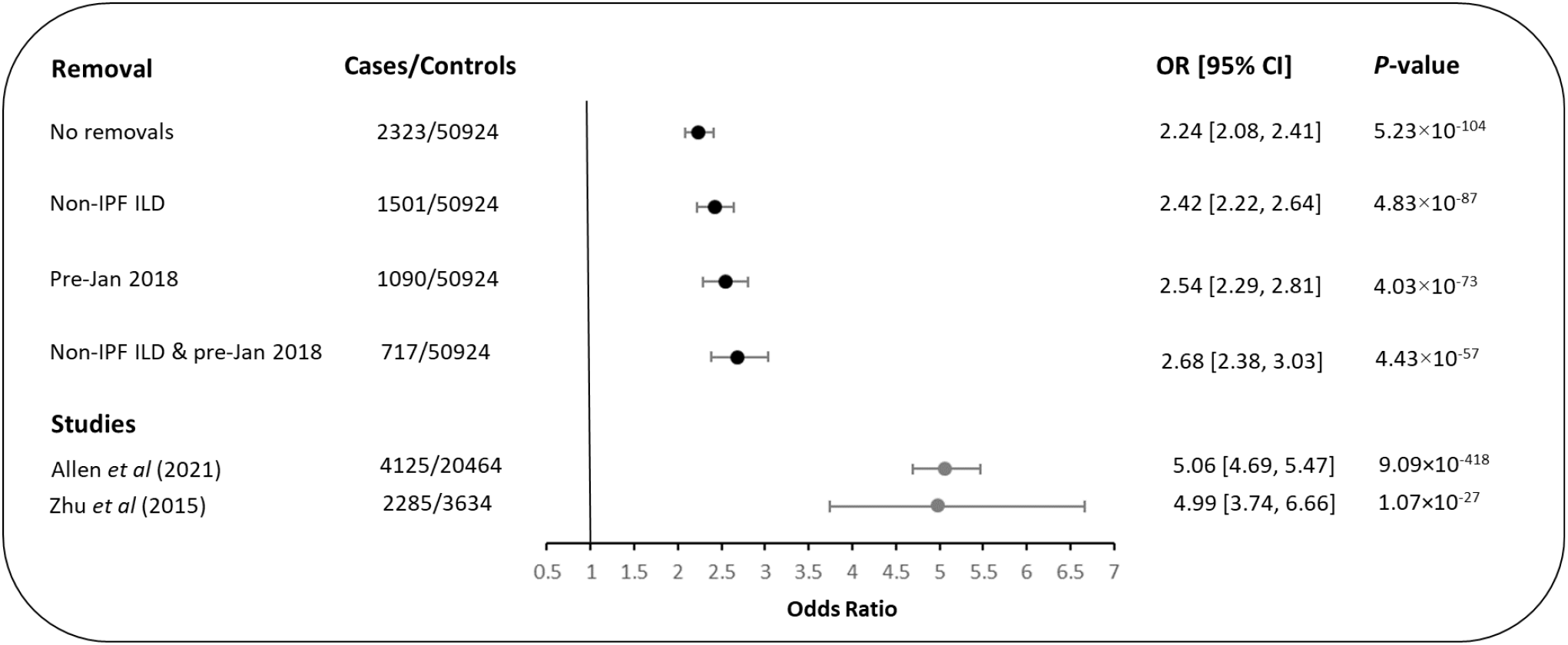
Effect size estimates of rs35705950 T allele association with IPF risk using ICD-10 codes and following exclusion of cases with a co-occurring code indicative of being non-IPF ILD or removing cases defined by the occurrence of a J84.1 code before January 2018. Each line shows the effect size estimate and confidence interval for the association between rs35705950 and IPF risk. Estimates in grey are the reference effect size estimates taken from Allen *et al* (2021) and Zhu *et al* (2015).

## Discussion

Our aim was to use the association with rs35705950 to evaluate the code-based definitions of IPF in UK Biobank. We show that none of the available IPF codes, either individually or in combination, replicate the effect size for the association of rs35705950 on IPF risk when using clinically-defined IPF cohorts. All code definitions did however provide a significant association. We observed that self-reported IPF in UK Biobank provided the best definition of IPF when evaluated using rs35705950 association, but this refined definition comes at the expense of sample size and hence statistical power.

We hypothesised that applying code-based exclusions to reduce misclassification amongst the cases, would improve the effect estimates. While this led to some increase in the effect sizes, this also rapidly reduced case sample size, and the effect estimates were still below the 95% confidence intervals of the estimates from IPF studies that used tertiary care diagnoses to recruit participants.

Within the ILD disease family the *MUC5B* SNP has been shown to be associated with other chronic progressive ILDs that exhibit a Usual Interstitial Pneumonia (UIP) radiological pattern (e.g. chronic hypersensitivity pneumonitis (cHP)). However, ILDs with a predominant nonspecific interstitial pneumonia (NSIP) radiological pattern (e.g. Systemic Sclerosis-associated ILD and Sarcoidosis) have not been associated with rs35705950. Excluding codes that might indicate a non-IPF ILD had an impact on the effect size estimates, but this was modest. We found that excluding J84.1 ICD-10 code entries that occurred prior to January 2018 was more effective at increasing the OR on its own than removing cases with co-occurring medical conditions that can cause pulmonary fibrosis.

We did not evaluate the impact of misdiagnosis with other common respiratory diseases such as asthma and chronic obstructive pulmonary disease (COPD). As misdiagnosis of asthma and COPD often occurs prior to a diagnosis of IPF (18), it is likely that excluding individuals with a co-occurring code for COPD or asthma would exclude true IPF cases.

We kept the same set of controls for our analyses for consistency when comparing effect size changes following changes to the case definition. Excluding individuals with a particular non-IPF code from the cases and not the control population could lead to associations related to that code in a GWAS. However, the rs35705950 association is specific to IPF-related codes and so we would not expect to be seeing changes to the effect size estimates due to pleiotropic effects of rs35705950.

In conclusion, UK Biobank offers an excellent resource for the study of low prevalence common diseases. However, for IPF, we show that commonly used codes fail to define a case sample that is able to robustly replicate previously reported association effect sizes. Furthermore, pragmatic attempts to refine the phenotype using further code exclusions were unable to improve the estimates. Studies of IPF using UK Biobank should take this into consideration (or discuss as a limitation) at study design stage and take into account the research purpose. For example, if the purpose is for new discovery, then gains in power due to sample size achievable by using the codes available may outweigh reductions in power due to misclassification. However, analyses that assume an accurately defined case population or that require true effect estimates should take the limitations of the disease definition into account. These findings are likely to be generalisable to studies of IPF in other UK general population cohorts with linked HES and primary care data.

## Data Availability

UK Biobank data is publicly accessible upon approval of an application through www.ukbiobank.ac.uk.

## Author Contributions

OCL, JKQ, RGJ and LVW developed the analysis plan. OCL, LMK and RJA performed the analyses. All authors contributed to developing IPF definitions. OCL and LVW wrote the first draft of the manuscript and all authors contributed and edited the final version.

## Data Availability

UK Biobank data is publicly accessible upon approval of an application through www.ukbiobank.ac.uk. UK Biobank has approval by the Research Ethics Committee (REC) under approval number 16/NW/0274.

