## Supplement for "Using genetic information to define idiopathic pulmonary fibrosis in UK Biobank"

### Supplementary Methods

#### **Data**

There were 488,377 samples genotyped in the UK Biobank and after removing participants who had withdrawn from UK Biobank and who had failed genotyping quality control (as described in Shrine *et al* 2019 (1)). Using the relationship inference software KING (2) we identified related participant pairs (2^nd^ degree or higher) and removed one individual from each pair. We assigned participants as cases if they had been defined as having IPF in at least one source. Controls were UK Biobank participants who had their primary care data linked but were not present in the IPF case subset. Controls were matched to cases based on age at admission [field: 21003], sex [field: 31], ever smoking status (as derived in Shrine *et al* (1) using a combination of fields: 1239, 1249 & 2644) and ethnicity (inferred using ADMIXTURE and K-means clustering, as described by Shrine *et al* (1)). We removed participants where these variables were missing.

For case-case and control-control pairs, *“ukb_gen_samples_to_remove”* from the *ukbtools* R package (3) was used to remove a subject in each pair using a greedy algorithm. For case-control pairs, the case was retained.

There were 2,629 IPF cases (IPF subset) of which composed of 2,303 HES cases, 580 mortality cases, 442 primary care cases, 142 self-reported PF cases, 107 self-reported IPF cases and eight with a medication code. For controls, there were 50,924 individuals.

**Supplementary Table 1: List of Medical conditions and codes for exclusion**

| **Condition** | **ICD-10 code** | **ICD-9 code** | **Non-cancer illness code**  **Data-Field: 20002** |
| --- | --- | --- | --- |
| Ankylosing spondylitis | M45 | 720* | 1313 |
| Amyloidosis | E85* | 277.3 |  |
| Asbestosis | J61 | 501 | 1120 |
|  | J92.0 |  |  |
| Chronic and other pulmonary manifestations due to radiation | J70.1 | 508.1 |  |
| Chronic respiratory conditions due to fumes or vapors | J68.4 | 506.4 |  |
|  | J68.9 |  |  |
| Coal worker's disease | J60 | 500 |  |
| Crohn's disease (Regional enteritis) | K50* | 555* | 1462 |
| Dermatomyositis - Polymyositis | M33* | 710.3 | 1383 |
|  |  | 710.4 | 1480 |
|  |  |  | 1481 |
| Extrinsic allergic alveolitis |  | 495* |  |
| Hypersensitivity pneumonitis due to organic dust | J67* |  |  |
| Idiopathic pulmonary hemosiderosis | J84.03 | 516.1 |  |
| Lipidosis | E75* | 272.7 |  |
|  | E77* |  |  |
| Lung involvement in other diseases classified elsewhere | J99* | 517.8 |  |
| Neurofibromatosis | Q85.0 | 237.7 |  |
| Other connective tissue diseases | M35* | 710.9 | 1377 |
|  |  |  | 1373 |
| Other metabolic disorders | E88* | 277.8 | 1496 |
| Other necrotizing vasculitis | M31* | 446* | 1376 |
|  |  |  | 1378 |
|  |  |  | 1379 |
| Pneumoconiosis | J62* | 502 |  |
|  | J63* | 503 |  |
|  | J64 | 504 |  |
|  | J65 | 505 |  |
|  | J66* |  |  |
| Polyarteritis nodosa | M30* | 446* | 1380 |
| Pulmonary alveolar proteinosis | J84.01 | 516.0 |  |
| Pulmonary eosinophilia | J82 | 518.3 |  |
| Pulmonary microlithiasis | J84.02 | 516.2 |  |
| Respiratory conditions due to other specified external agents | J70.9 | 508.8 |  |
| Rheumatoid arthritis and rheumatoid lung | J99.0 | 714* | 1464 |
|  | M05* |  |  |
|  | M06* |  |  |
| Sarcoidosis | D86* | 135* | 1371 |
| Sjogren syndrome | M35.0 | 710.2 | 1382 |
| Systemic lupus erythematosus | M32* | 710.0 | 1381 |
| Systemic sclerosis | L94* | 710.1 | 1384 |
|  | M34* | 517.2 |  |
| Tuberous sclerosis | Q85.1 | 759.5 |  |
| Note: Adapted from e-Table 1 in Bellou *et al* (4): Bellou V, Belbasis L, Evangelou E. *Tobacco smoking and risk for pulmonary fibrosis: a prospective cohort study from the UK Biobank*. Chest. 2021 | | | |

**Supplementary Table 2: rs35705950 minor allele frequency and association results in UK Biobank defined IPF groups**

|  | **IPF definition (European ancestry only)** | | | | | |  |  |
| --- | --- | --- | --- | --- | --- | --- | --- | --- |
|  | **IPF HES** | **IPF mortality** | **Self-reported IPF** | **Self-reported pulmonary fibrosis** | **IPF primary care** | **IPF subset**  **(all IPF subsets combined)** | **Allen *et al***  **(2021)** | **Zhu *et al***  **(2015)** |
| **Sample size** |  |  |  |  |  |  |  |  |
| Cases | 2,217 | 561 | 104 | 139 | 427 | 2,535 | 3,584* | 2,285 |
| Controls | 50,924 | 50,924 | 50,924 | 50,924 | 50,924 | 50,924 | 19,922* | 3,634 |
| **T allele frequency** |  |  |  |  |  |  |  |  |
| Cases | 22% | 25% | 28% | 21% | 27% | 22% | 33% | NA |
| Controls | 11% | 11% | 11% | 11% | 11% | 11% | 11% | NA |
| ORs [95% CI] | 2.23 [2.07, 2.40] | 2.62 [2.29, 3.01] | 3.09 [2.28, 4.18] | 2.06 [1.54, 2.76] | 2.95  [2.53, 3.44] | 2.26 [2.10, 2.42] | 5.06  [4.69, 5.47] | 4.99  [3.74, 6.66] |
| *p*-value | 1.05×10^-98^ | 3.81e×10^-43^ | 3.33×10^-13^ | 1.00×10^-6^ | 8.37×10^-44^ | 9.74×10^-115^ | 9.09×10^-418^ | 1.07×10^-27^ |
| Note: coded allele = T, rs35705950 dosages were used to perform the analysis  Association model: IPF status = rs35705950 + PC1 + PC2 + PC3 + PC4 + PC5 + PC6 + PC7 + PC8 + PC9 + PC10 Where, PC1 to PC10 are the first 10 ancestry principal components (UK Biobank data-Field 22009), rs35705950 are the dosages for rs35705950  OR = odds ratio and 95% CI = 95% confidence interval for odds ratio  * rs35705950 was not available in one of the discovery cohorts (Chicago study) due to poor imputation quality, so sample sizes are smaller than for the full cohort | | | | | | | | |

**Supplementary Table 3: rs35705950 minor allele frequency and association results in UK Biobank defined IPF groups HES and mortality combined after removals used to refine IPF case definition**

| **IPF definition (European ancestry only)** | | | | | | | | | | | | |
| --- | --- | --- | --- | --- | --- | --- | --- | --- | --- | --- | --- | --- |
|  | **No removals** | | | **Non-IPF ILD** | | | **Pre-Jan 2018** | | | **Non-IPF ILD & pre-Jan 2018** | | |
|  | **HES** | **Mortality** | **HES and mort combined** | **HES** | **Mortality** | **HES and mort combined** | **HES** | **Mortality** | **HES and mort combined** | **HES** | **Mortality** | **HES and mort combined** |
| **Sample size** |  |  |  |  |  |  |  |  |  |  |  |  |
| Cases | 2,217 | 561 | 2,323 | 1,418 | 392 | 1,501 | 1,023 | 182 | 1,090 | 671 | 127 | 717 |
| Controls | 50,924 | 50,924 | 50,924 | 50,924 | 50,924 | 50,924 | 50,924 | 50,924 | 50,924 | 50,924 | 50,924 | 50,924 |
| **T allele frequency** |  |  |  |  |  |  |  |  |  |  |  |  |
| Cases | 22% | 25% | 22% | 23% | 26% | 23% | 24% | 24% | 24% | 25% | 26% | 25% |
| Controls | 11% | 11% | 11% | 11% | 11% | 11% | 11% | 11% | 11% | 11% | 11% | 11% |
| ORs [95% CI] | 2.23 | 2.62 | 2.24 | 2.39 | 2.83 | 2.42 | 2.56 | 2.57 | 2.54 | 2.68 | 2.82 | 2.68 |
|  | [2.07, 2.40] | [2.29, 3.01] | [2.08, 2.41] | [2.19, 2.62] | [2.41, 3.33] | [2.22, 2.64] | [2.31, 2.84] | [2.00, 3.29] | [2.29, 2.81] | [2.37, 3.04] | [2.13, 3.73] | [2.38, 3.03] |
| *p*-value | 1.05×10^-98^ | 3.81×10^-43^ | 5.23×10^-104^ | 2.89×10^-80^ | 5.75×10^-37^ | 4.83×10^-87^ | 1.82×10^-70^ | 1.50×10^-13^ | 4.03×10^-73^ | 1.01×10^-53^ | 4.38×10^-13^ | 4.43×10^-57^ |
| Note: coded allele = T, rs35705950 dosages were used to perform the analysis Association model: IPF status = rs35705950 + PC1 + PC2 + PC3 + PC4 + PC5 + PC6 + PC7 + PC8 + PC9 + PC10 Where, PC1 to PC10 are the first 10 ancestry principal components (UK Biobank data-Field 22009), rs35705950 are the dosages for rs35705950 OR = odds ratio and 95% CI = 95% confidence interval for odds ratio **No removals** = association results before any removals **Non-IPF ILD** = after removal of those who have a code for non-IPF medical conditions that cause pulmonary fibrosis  **Pre-Jan 2018** = after removal of J84.1 ICD-10 code occurrence that occurred before January 2018 | | | | | | | | | | | | |

**Supplementary Figure 1: Venn diagram to examine participant overlap between IPF sources in UK Biobank in those that have non-missing HES, primary care, self-reported IPF and self-reported PF (n = 202)**

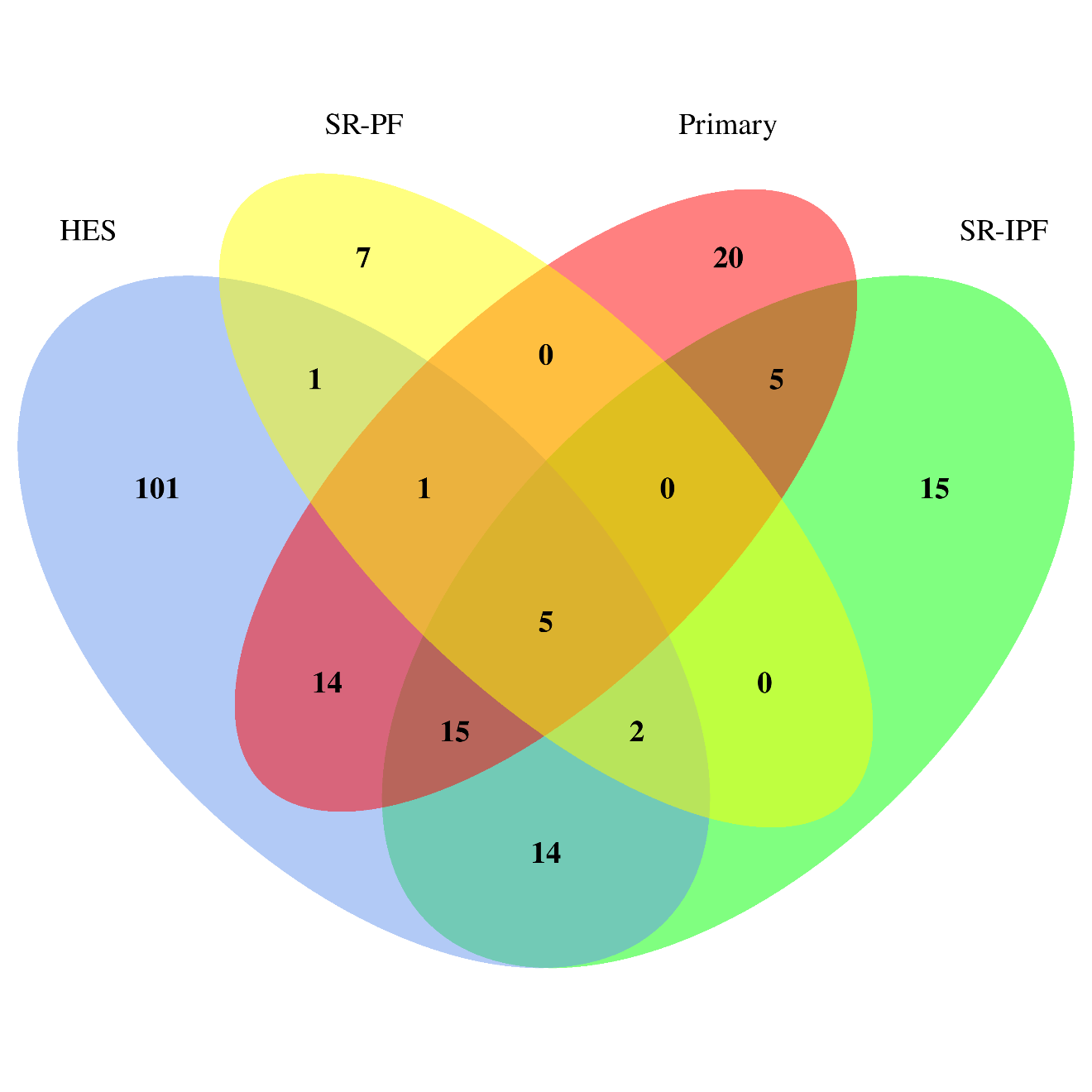

Note: **HES** = Hospital Episodes Statistics data, **Primary** = primary care data, **SR-PF** = self-reported pulmonary fibrosis and
**SR-IPF** = self-reported IPF
